# Standard Model Imaging for Discriminating Multiple Sclerosis Lesion Types: A Lesion-Focused Analysis Compared with Diffusion Tensor Imaging

**DOI:** 10.64898/2026.03.15.26348424

**Authors:** Chaoyang Jin, Ahmad Tubasi, Ke Xu, Caroline Gheen, Taegan Vinersky, Hakmook Kang, Xiaoyu Jiang, Francesca Bagnato, Junzhong Xu

## Abstract

**Purpose:** To characterize microstructural alterations across distinct white matter tissue classes in MS using Standard Model Imaging (SMI), and to place its performance in context relative to conventional diffusion tensor imaging (DTI).

**Methods:** DTI and SMI were applied to treatment-naïve individuals at early stages of MS, including patients with MS and healthy controls. Over 3,602 manually delineated regions of interest were classified into normal white matter (NWM), normal-appearing white matter (NAWM), T2-hyperintense lesions, and chronic black holes (cBHs) differences were assessed using linear mixed-effects models with false discovery rate correction. Discriminative performance was evaluated using receiver operating characteristic (ROC) analysis within a generalized linear mixed modeling framework for individual parameters and multivariate DTI, SMI, and combined DTI+SMI models.

**Results:** Both DTI and SMI metrics demonstrated widespread and significant differences across tissue classes. Robust discriminative performance was observed for lesion-NWM and lesion-NAWM comparisons (AUC > 0.8), whereas discrimination between NAWM and NWM and between cBHs and T2-lesions was limited (AUC ≤ 0.66). In terms of model performance, SMI achieved slightly higher AUC values than DTI across most contrasts, while the combined DTI+SMI model consistently provided the highest diagnostic performance. ROI-based analyses revealed additional SMI alterations, including changes in extra-axonal parallel diffusivity, not consistently reported in prior studies.

**Conclusion:** DTI and SMI metrics are sensitive to microstructural abnormalities across a broad spectrum of white matter tissue classes in MS, capturing both lesion-related damage and more subtle alterations extending into NAWM. While discriminative performance varies by tissue contrast, integrating DTI and SMI provides complementary information and modestly improves diagnostic performance, supporting a multi-model diffusion MRI approach for comprehensive characterization of MS-related white matter pathology

## 1 Introduction

Multiple sclerosis (MS) is a chronic autoimmune disorder of the central nervous system characterized by inflammation, demyelination, and axonal injury[1, 2]. Although MS often presents with a relapsing course, axonal degeneration begins early and progresses over time[3]. Magnetic resonance imaging (MRI) is central to MS diagnosis and monitoring, with conventional T1- and T2-weighted sequences enabling the detection of focal lesions and brain atrophy[4, 5]. Active inflammatory lesions appear as gadolinium-enhancing abnormalities, most of which evolve into chronic T2-hyperintense lesions, while a subset persists as T1-hypointense chronic black holes (cBHs), reflecting severe axonal loss[6]. However, conventional imaging markers lack pathological specificity and quantitative sensitivity, contributing to poor correlations with clinical disability[7]. This limitation largely stems from the insensitivity of standard MRI to subtle microstructural damage in normal-appearing white matter (NAWM) and gray matter, underscoring the need for advanced imaging biomarkers capable of quantifying diffuse neurodegenerative processes in MS[8, 9]. Diffusion MRI (dMRI) has emerged as a pivotal tool for detecting subtle microstructural pathology in MS, particularly within NAWM that remains undetectable on conventional MRI sequences[10]. By quantifying the microscopic displacement of water molecules, dMRI provides sensitive markers of tissue integrity[11]. Standard diffusion tensor imaging (DTI), the most widely applied approach, yields metrics such as axial diffusivity (AD), commonly interpreted as a surrogate for axonal integrity[12]. However, DTI metrics lack pathological specificity, as they reflect compartment-averaged diffusion signals and are influenced by factors such as fiber orientation and crossing[12]. To overcome these limitations, biophysical modeling frameworks such as the Standard Model Imaging (SMI) have been introduced to derive compartment-specific metrics of axonal density, diffusivity, and fiber organization from diffusion MRI data[13].

Although the technical reliability of SMI has been demonstrated on clinically feasible 3T MRI systems, its ability to characterize the full spectrum of microstructural pathology in MS remains incompletely understood[14]. Recent work by Liao et al. extended SMI to MS cohorts, demonstrating reduced intra-axonal water fraction (*f*), reduced orientation coherence metric (*p*_2_), along with increased extra-axonal perpendicular diffusivity 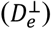 in MS patients relative to healthy controls, supporting the sensitivity of SMI metrics to diffuse microstructural abnormalities[15]. However, these findings were largely derived from automated atlas-based segmentation that includes entire white matter tracts containing both lesions and NAWM, which reduces lesion specificity and limits direct comparability with standard clinical approaches based on manually delineated lesion ROIs [16] [17].

These limitations underscore the clinical standard of using ROI-based, lesion-focused analyses that explicitly incorporate lesion interiors, enabling systematic and biologically interpretable comparisons across the spectrum of white matter pathology in MS [18]. In response, the present study establishes such a framework spanning five white matter categories, including normal appearing tissue and distinct lesion subtypes such as T2-lesions and cBHs [19]. Leveraging 3602 manually delineated regions of interest, this approach enables lesion-resolved analyses that capture microstructural heterogeneity across the continuum from normal to severely damaged white matter.

Here, we conducted this study to address the limited pathological specificity of conventional diffusion MRI in early multiple sclerosis and the lack of systematic comparisons across advanced diffusion models at the lesion level. Prior work has largely relied on single-shell DTI, which provides sensitive but non-specific markers of tissue damage and limited ability to disentangle heterogeneous microstructural alterations across MS tissue classes. Although higher-order diffusion models such as SMI may offer improved microstructural specificity, their added value relative to DTI across distinct MS tissue compartments remains insufficiently explored, particularly in earlystage disease. To address these gaps, we applied DTI and SMI to individuals at early stages of multiple sclerosis, including patients with newly diagnosed multiple sclerosis (pwMS) [20, 21]. We systematically compared diffusion-derived metrics across lesions and non-lesioned white matter at both the individual parameter and model levels. We hypothesized that diffusion-based metrics would capture graded microstructural alterations across tissue classes, and that SMI would provide complementary microstructural information to conventional DTI in characterizing MS-related white matter pathology.

## 2 Methods

### 2.1 Study design and cohort

The study protocol was approved by the Institutional Review Boards of Vanderbilt University Medical Center and the Nashville VA Medical Center. All procedures were conducted in accordance with the ethical principles of the Declaration of Helsinki. Written informed consent was obtained from all participants.

This study enrolled 57 pwMS and 17 age-matched healthy controls. Participants were excluded if they had any of the following: contraindications to MRI, history of ischemic or hemorrhagic stroke, other systemic or central nervous system autoimmune disorders, active neoplastic or infectious disease, uncontrolled hypertension or diabetes mellitus, clinically significant cardiac disease, or prior exposure to MS disease modifying therapies (with the exception of glucocorticoids used for acute relapse management). Additionally, participants who experienced clinical changes between neurological assessment and MRI acquisition were excluded.

### 2.2 MRI acquisition

Brain MRI examinations were performed using a 3.0-Tesla Ingenia CX scanner (Philips Healthcare, Best, The Netherlands) with a volume transmit and 32-channel receive head coil (NOVA Medical, Wilmington, MA, USA). The imaging protocol included axial T1-weighted turbo spin echo (T1w TSE), axial T2-weighted fluid-attenuated inversion recovery (T2-FLAIR), and multi-shell diffusion MRI (dMRI). In the patient cohort, contrast-enhanced imaging was performed to assess gadolinium-enhancing lesions, using either pre- and post-contrast 3D magnetization-prepared rapid acquisition gradient echo (3D MPRAGE) sequences or post-contrast T1-weighted TSE sequences. All sequences provided whole-brain coverage. Detailed acquisition parameters are provided in **Table 1**.

**Table 1.** Pulse Sequence Parameters.

|  | T<sub>1</sub>-w TSE | T<sub>2</sub>-w FLAIR | dMRI |
| --- | --- | --- | --- |
| Acquisition time (minutes) | ~3 | ~5 | ~8 |
| Compressing SENSE | 1.80 | None | 2 |
| Field of view(cm) | 21 × 17 × 12.8 | 21 × 17 × 12.8 | 21 × 20.2 × 12.8 |
| Acquired voxel size(mm) | 1 × 1 × 2 | 1 × 1 × 2 | 2(istropic) |
| Echo time (msec) | 125 | 9.4 | 90 |
| Inversion time (seconds) | NA | NA | NA |
| Repetition time (seconds) | 11 | 0.6-0.7 | 4.5 |
| Number of b-shells | NA | NA | 2(b = 711 sec/mm<sup>2</sup> with 32 directions and b = 2855 sec/mm<sup>2</sup> with 64 directions) |
| Multi-band factor | NA | NA | 2 |

NA = not applicable; T1-w TSE = T1-weighted turbo spin echo; T2-w FLAIR = T2-weighted fluid attenuated inversion recovery image; DTI = diffusion tensor imaging;

### 2.3 Image analysis

#### dMRI pre-processing

For each subject, DTI data were corrected for Gibbs ringing artefacts using the mrdegibbs tool in the MRtrix3 (https://www.mrtrix.org/)[22], and susceptibility-induced distortions and eddy current artifacts using the top-up and eddy tools from the FMRIB Software Library (FSL; https://fsl.fmrib.ox.ac.uk/fsl/)[23].

#### Parametric map calculations

DTI analysis was performed using MRtrix3 to yield fractional anisotropy (FA), axial diffusivity AD, radial diffusivity (RD), and mean diffusivity (MD). SMI parameters estimation was conducted using the open-source toolbox developed by the NYU Diffusion MRI Group (https://github.com/NYU-DiffusionMRI/SMI). This framework models white matter as a two-compartment system (intra- and extra-axonal spaces) with orientation dispersion captured via spherical harmonic expansion. Five voxel-wise parametric maps were generated from the multi-shell dMRI data (See Figure 1). No post-processing spatial smoothing or denoising was applied to preserve native spatial resolution.

**Figure 1.**
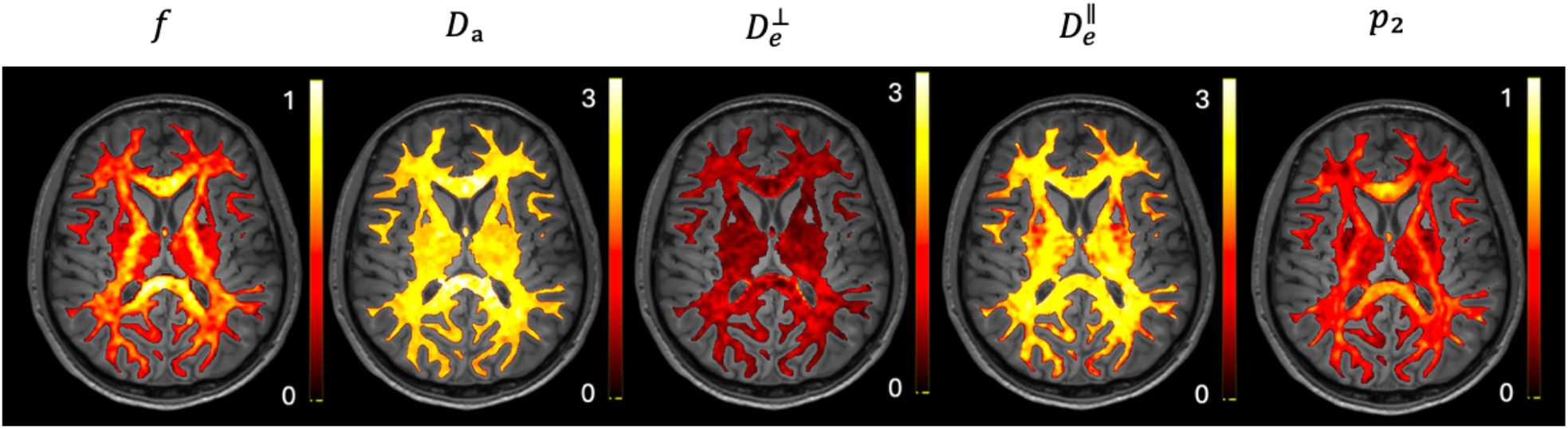
Five parameters mapping of SMI model. *f* intra-axonal volume fraction; *D*_*a*_ intra-axonal diffusivity;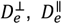 extra-axonal perpendicular and parallel diffusivity, respectively; *p*_2_ orientation coherence metric.

**Figure 2.**
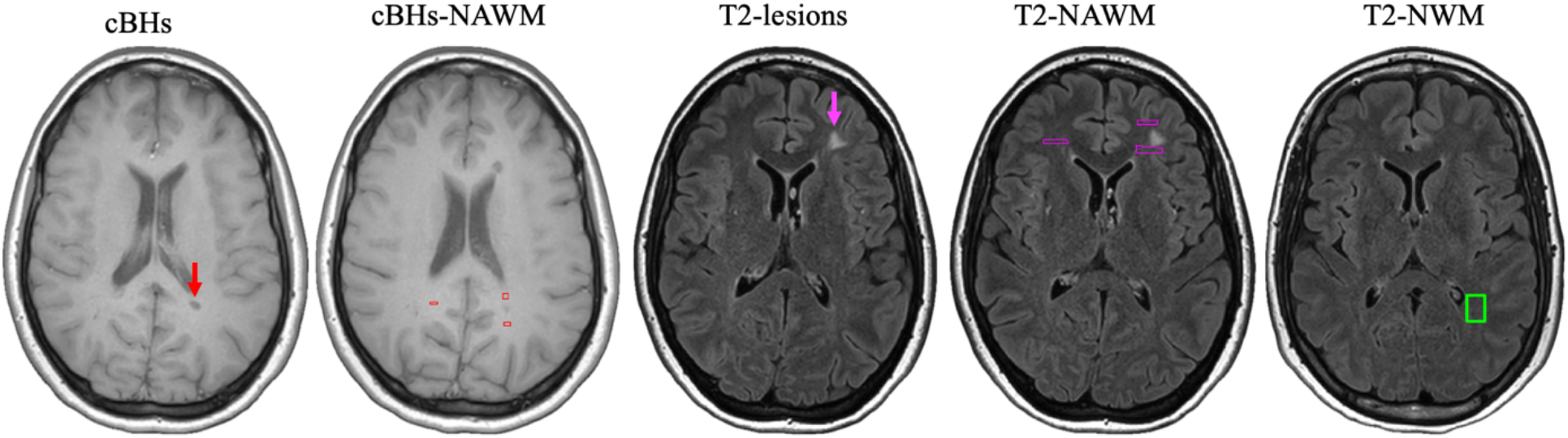
Representative examples of lesion identification and region of interest (ROI) placement. The figure displays the segmentation strategy across T1-weighted and T2-FLAIR sequences: (A) T1-weighted image showing a chronic black hole (cBH), indicated by a red arrow. (B) T1-weighted image illustrating cBH-associated NAWM ROIs (red boxes), placed proximally to the lesions. (C) T2-FLAIR image displaying a T2-hyperintense lesion, indicated by a pink arrow. (D) T2-FLAIR image showing lesion-associated NAWM ROIs (purple boxes), placed proximally to the T2-hyperintensities. (E) T2-FLAIR image showing a normal white matter (NWM) ROI (green box) in a healthy control. Abbreviations: cBHs, chronic black holes; NAWM, normal-appearing white matter; NWM, normal white matter.

#### Image registration

All structural and diffusion-derived images, including T1-w TSE, SMI parametric maps, and DTI-derived maps, were then spatially registered to the corresponding T2 FLAIR image using affine transformation implemented via the FSL Linear Image Registration Tool (FLIRT). This approach ensured anatomical alignment across modalities and has been validated in prior study[24].

#### Image analysis

We employed the Medical Imaging Processing, Analysis, and Visualization (MIPAV) software (version 7.3; https://mipav.cit.nih.gov/) for manual delineation of lesions and non-lesional regions of interest (ROIs) and for quantification of microstructural metrics[25]. A senior neuroradiologist (F.B., with more than 20 years of experience) initially identified all gadolinium-enhancing lesions on post-contrast T1-weighted images. These enhancing lesions were excluded from subsequent analysis to avoid biological confounding from acute inflammation and edema.

Following this screening, a postdoctoral research fellow with a medical degree (A.T.) manually segmented all T2-lesions and cBHs. To rigorously account for anatomical variability, a matched-ROI approach was implemented. In patients, corresponding NAWM ROIs were defined according to their spatial relation to the lesion: proximal NAWM, immediately adjacent anteriorly and posteriorly to the lesion, and distant NAWM, located in the contralateral hemisphere. In healthy controls, anatomically corresponding normal white matter ROIs were drawn to match the locations of lesions or NAWM ROIs in patients. All generated masks were subsequently reviewed by the senior neuroradiologist (F.B.) to confirm accuracy and consistency. Finally, the validated binary ROI masks were overlaid onto co-registered SMI and DTI parametric maps to extract mean values for each region.

### 2.4 Statistical Analyses

#### Microstructural Differences Across Tissue Types

Microstructural differences among tissue classes (cBHs, T2-lesions, NAWM in patients, and NWM in healthy controls) were evaluated using linear mixed-effects models (LMM)[26]. This approach appropriately accounted for the hierarchical data structure, with multiple ROIs nested within individual subjects. For each microstructural metric (e.g., *f, p*_2_, FA, MD), the metric was specified as the dependent variable, tissue type as the fixed effect, and subject as a random intercept to adjust for within-subject, with age and sex included as covariates. Post-hoc pairwise comparisons among tissue classes were conducted with false discovery rate (FDR) correction for multiple testing.

To preserve anatomical specificity, ROI-based measurements were further organized using a tract-informed framework. Major white matter tracts were segmented in native diffusion space using TractSeg, enabling subject-specific tract delineation without inter-subject registration[27]. Each ROI was spatially mapped onto the tract masks and assigned to the tract showing the greatest voxel-wise overlap, ensuring anatomically consistent tract classification while preserving individual variability. Tract-informed ROI metrics were then incorporated into the linear mixed-effects models with subject included as a random effect, allowing assessment of tissue-class–related microstructural differences while accounting for both within-subject clustering and tract-level anatomical organization.

#### Discriminative performance and Model Comparison

The discriminative ability of diffusion-derived microstructural metrics was evaluated using receiver operating characteristic (ROC) analysis within a generalized linear mixed modeling (GLMM) framework with a binomial distribution and logit link function[28]. First, individual microstructural parameters from DTI and SMI were assessed separately by fitting single-parameter GLMM, adjusted for age and sex with a random intercept for study, to quantify the discriminative performance of each metric for specific tissue contrasts (e.g., cBHs vs T2-lesions, cBHs vs cBHs-NAWM). To evaluate overall classification performance, three multivariate GLMM were constructed: an SMI model incorporating all five somatic microstructural imaging parameters 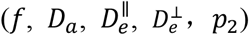; a DTI model incorporating all four diffusion tensor imaging parameters (FA, MD, AD, RD); and a joint model integrating parameters from both SMI and DTI.

## 3 Results

### 3.1 Participants Characteristics

A total of 57 MS patients and 17 healthy controls (HCs) were included in this study. The mean age of MS patients was 38.82 ± 11.70 years, and that of HCs was 36.94 ± 12.10 years. There was no significant difference in age between the two groups (Welch’s t-test, p = 0.577). In terms of sex distribution, the MS group included 39 females and 18 males, while the HC group included 11 females and 6 males. There was no significant difference in sex distribution between groups (χ^2^ test, p = 0.774).

### 3.2 Differences in SMI and DTI Parameters Between Tissue Classes

In total, more than 3,000 manually delineated regions of interest were included in the present study, comprising 223 cBHs, 270 cBHs-NAWM, 1,621 T2-lesions, 1,196 T2-NAWM, and 292 T2-NWM.

#### 3.2.1 Lesions and NWM

For the comparison between cBHs and NWM, widespread and significant differences were observed across nearly all DTI and SMI metrics (Fig. 3). Specifically, cBHs exhibited reduced FA, accompanied by increased MD, AD, and RD in the DTI model. In the SMI framework, cBHs showed a decreased *f p*_2_ and Increased 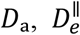 and 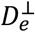. A similar pattern was observed for T2 lesions relative to NWM, with consistent alterations across DTI and SMI metrics, except that *p*_2_ showed no significant difference.

**Figure 3.**
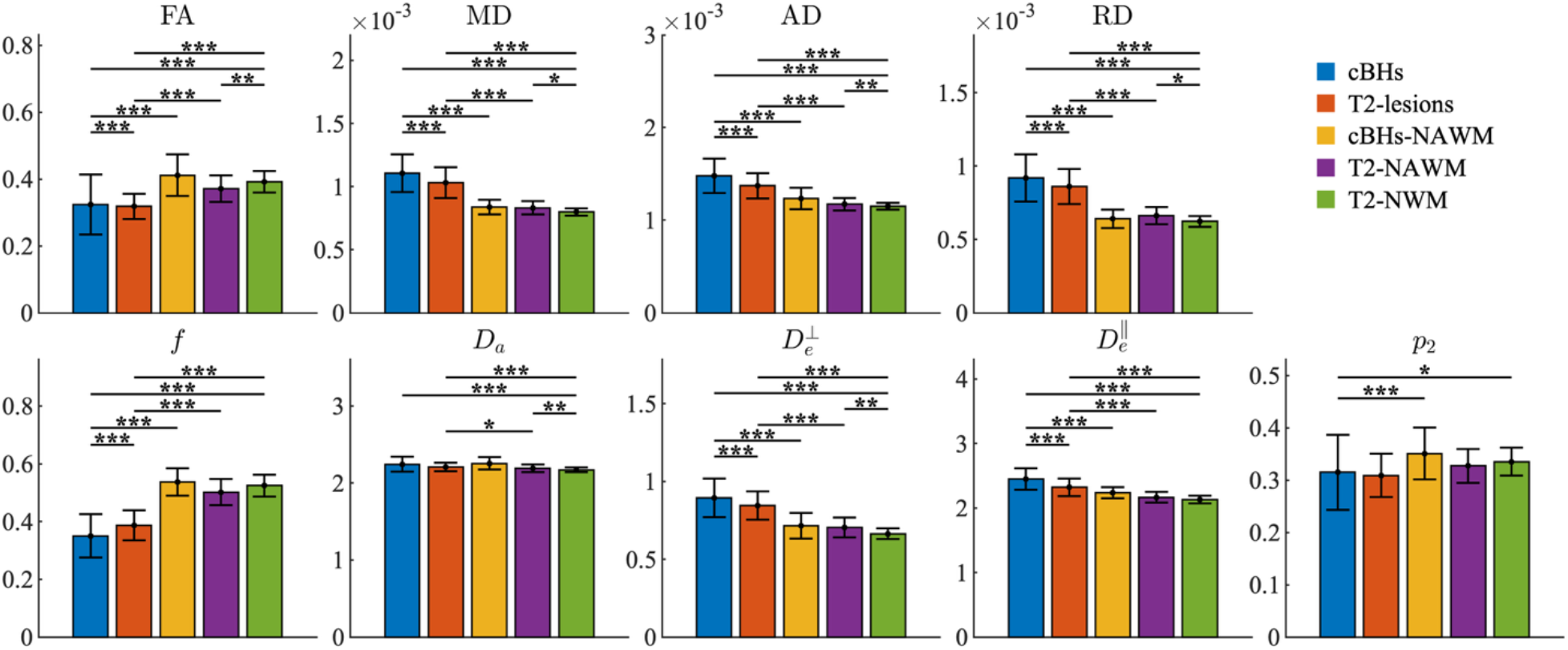
Group comparisons of diffusion metrics derived from DTI and SMI models across five white matter tissue types in MS and HC: cBHs, chronic black holes; T2-lesions, NAWM, Normal Appearing White Matter; NWM, Normal White Matter; Fractional Anisotropy, FA; Mean Diffusivity, MD; Axial diffusivity, AD; Radial Diffusivity, RD; Intra-Axonal Volume Fraction, *f*, Intra-Axonal Diffusivity, *D*_a_; Extra-Axonal Parallel Diffusivity,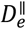; Extra-Axonal Perpendicular Diffusivity,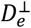; and fiber orientation coherence,*p*_2_. Error bars indicate standard deviation across subjects. Asterisks denote statistical significance: P < 0.05 (*), P < 0.01 (**), FDR-corrected.

#### 3.2.1 Lesions and NAWM

When comparing lesions with NAWM, both cBHs and T2-lesions exhibited highly consistent alterations in the DTI model, characterized by significantly reduced FA and increased MD, AD, and RD. In the SMI model, cBHs showed a largely concordant pattern, with decreased intra-axonal volume fraction *f* and increased 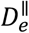 and 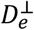. With respect to fiber orientation coherence, cBHs demonstrated a significant reduction in *p*_2_, whereas no significant difference in *p*_2_ was observed for T2-lesions. By contrast, *D*_a_ was significantly increased in T2 lesions, whereas no significant change was detected in cBHs (Fig. 3).

#### 3.2.3 NAWM and NWM

Significant differences between T2-NAWM and T2-NWM were observed across both DTI and SMI metrics. T2-NAWM exhibited reduced FA and increased MD, AD, and RD in the DTI model, along with elevated *D*_a_ and 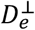 in the SMI framework (Fig. 3).

#### 3.2.4 cBHs and T2-lesions

In the comparison between cBHs and T2 lesions, all DTI parameters exhibited significant differences, characterized by reduced FA and increased MD, AD, and RD. In the SMI model, significant increases were observed in *D*_a_ and 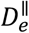.

### 3.3 Discriminative Performance of SMI and DTI Parameters Across Tissue Classes

#### 3.3.1 Lesions and NWM

For both cBHs vs T2-NWM and T2-lesions vs T2-NWM comparisons, all DTI parameters achieved robust discriminative performance. Similarly, within the SMI model, *f*,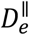, and 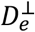 consistently provided strong separation between tissue classes (Fig. 4; Table 4).

**Table 4.** AUC values for SMI and DTI parameters across six pairwise tissue comparison.

| AUC | Parameters | cBHs vs. T2-NWM | T2-lesions vs. T2-NWM | cBHs vs cBHs-NAWM | T2-Lesions vs. T2-NAWM | T2-NAWM vs. T2-NWM | cBHs vs T2-Lesions |
| --- | --- | --- | --- | --- | --- | --- | --- |
| SMI | $f$ | 0.88 | 0.80 | 0.90 | 0.74 | 0.59 | 0.66 |
| | $D_a$ | 0.66 | 0.60 | 0.59 | 0.53 | 0.56 | 0.57 |
| | $D_e^{\parallel}$ | 0.84 | 0.70 | 0.75 | 0.65 | 0.55 | 0.60 |
| | $D_e^{\perp}$ | 0.89 | 0.80 | 0.84 | 0.71 | 0.61 | 0.60 |
| | $p_2$ | 0.60 | 0.55 | 0.66 | 0.52 | 0.56 | 0.55 |
| DTI | AD | 0.86 | 0.75 | 0.77 | 0.71 | 0.55 | 0.60 |
|  | FA | 0.76 | 0.69 | 0.79 | 0.63 | 0.57 | 0.61 |
|  | MD | 0.94 | 0.85 | 0.90 | 0.77 | 0.62 | 0.62 |
|  | RD | 0.92 | 0.83 | 0.90 | 0.75 | 0.61 | 0.61 |

**Figure 4.**
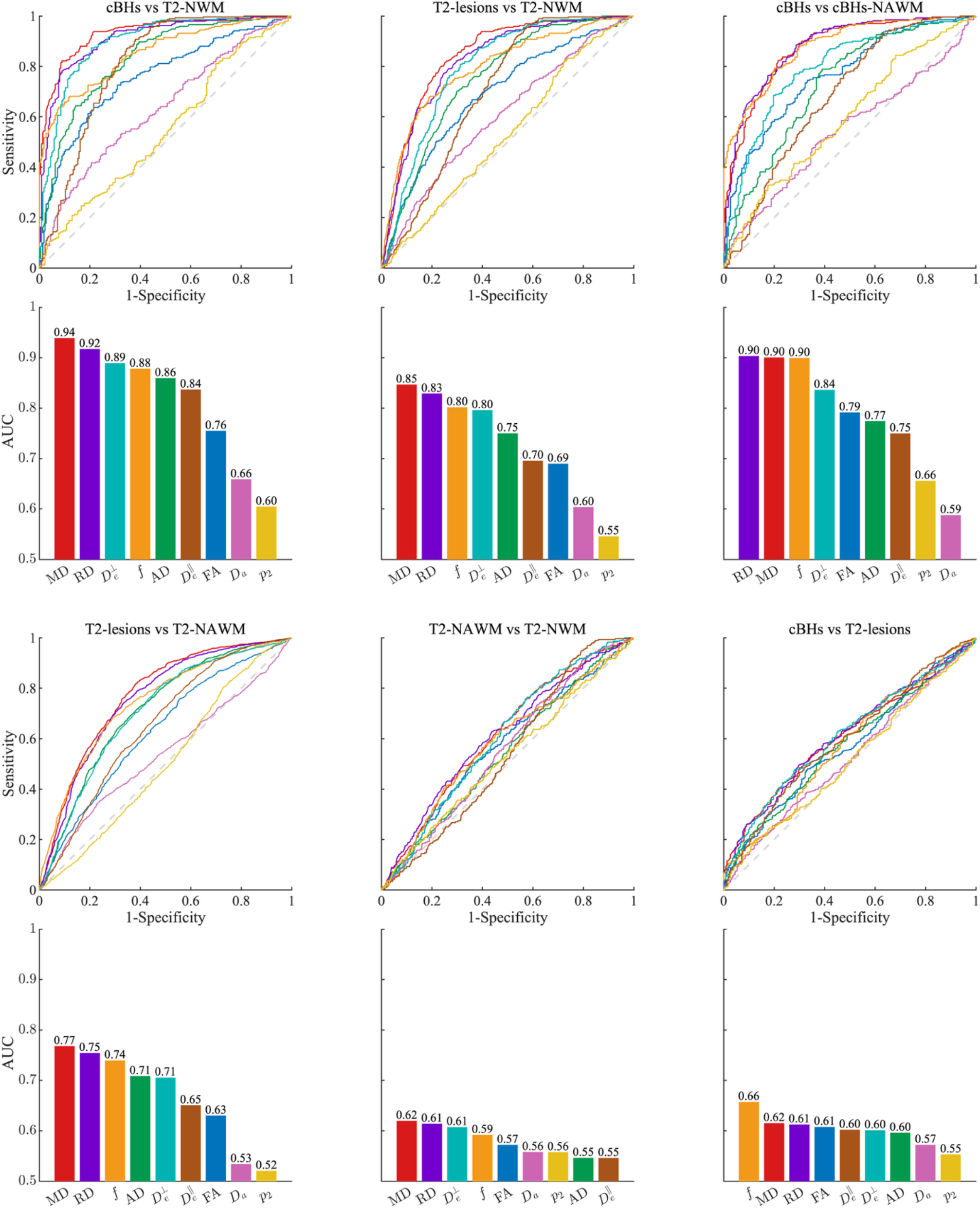
ROC analysis of DTI and SMI parameters across four white matter tissue contrasts. cBHs, chronic black holes; T2-lesions, NAWM, Normal Appearing White Matter; NWM, Normal White Matter; Fractional Anisotropy, FA; Mean Diffusivity, MD; Axial diffusivity, AD; Radial Diffusivity, RD; Intra-Axonal Volume Fraction, *f*, Intra-Axonal Diffusivity, *D*a; Extra-Axonal Parallel Diffusivity,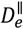; Extra-Axonal Perpendicular Diffusivity,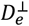; and fiber orientation coherence, *p*_2_.

#### 3.3.2 Lesions and NAWM

Across lesion–NAWM contrasts, both DTI and SMI parameters demonstrated meaningful discriminative performance, with subtype-specific patterns. All DTI metrics exhibited strong discrimination between cBHs and cBHs-NAWM. Within the SMI framework, robust discrimination was observed for *f*,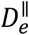, and 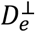. For T2-lesions relative to T2-NAWM, discriminative performance among DTI parameters was primarily driven by MD, RD, and AD, while within the SMI model, *f* and 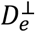 provided effective separation between tissue classes (Fig. 4; Table 4).

#### 3.3.3 NAWM and NWM

In the T2-NAWM versus T2-NWM contrast, neither DTI nor SMI parameters demonstrated strong discriminative performance. Only MD, RD, and 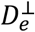 achieved modest discrimination, with AUC values exceeding 0.6 (Fig. 4; Table 4).

#### 3.3.4 cBHs and T2-lesions

In the cBHs versus T2-lesions contrast, overall discriminative performance was limited across both DTI and SMI parameters. Among all metrics, only the SMI-derived f parameter showed modest discriminative ability, with an AUC of 0.66 (Fig. 4; Table 4).

### 3.4 Discriminative Performance of DTI, SMI, and Combined Models

As shown in Figure 5, across all tissue contrasts, SMI-based models demonstrated slightly higher discriminative performance than DTI-based models, although the overall performance of the two approaches was largely comparable. Notably, the combined DTI+SMI model consistently achieved the highest AUC values across all comparisons.

**Figure 5.**
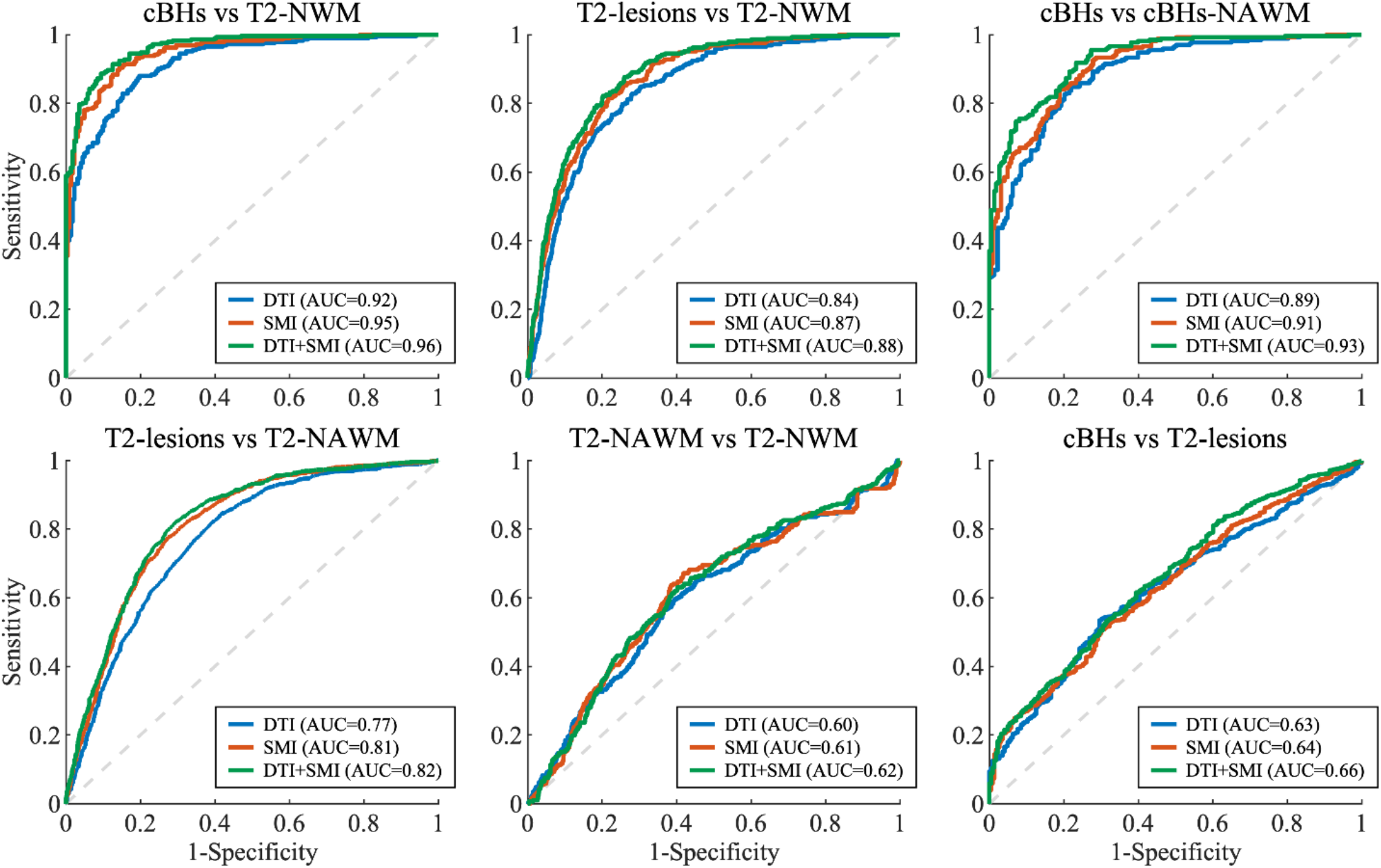
ROC curves comparing the discriminative performance of SMI, DTI, and DTI+SMI models across four tissue contrasts. AUC values are shown in the legend.

#### 3.4.1 Lesions and NWM

Both DTI and SMI models demonstrated strong discriminative performance in differentiating cBHs vs T2-NWM and T2-lesions vs T2-NWM, with AUC values exceeding 0.8. Across these contrasts, SMI consistently achieved slightly higher AUC values than DTI (e.g., 0.95 vs 0.92 for cBHs vs T2-NWM, and 0.87 vs 0.84 for T2-lesions vs T2-NWM). Notably, the combined DTI+SMI model yielded the highest classification performance, reaching AUC values of 0.96 and 0.88, respectively.

#### 3.4.2 Lesions and NAWM

In the lesion-NAWM comparisons, including cBHs vs cBHs-NAWM and T2-lesions vs T2-NAWM, DTI and SMI demonstrated comparable discriminative performance. SMI achieved slightly higher AUCs than DTI (e.g., 0.91 vs 0.89 and 0.81 vs 0.77), while the combined DTI+SMI model showed the best overall performance (0.93 and 0.82).

#### 3.4.3 NAWM and NWM

Discrimination between T2-NAWM and T2-NWM remained challenging, with all models yielding relatively low AUCs, and the highest performance observed for the combined DTI+SMI model (AUC=0.62).

#### 3.4.3 cBHs and T2-lesions

Distinguishing cBHs from T2-lesions proved difficult, with all models showing limited discriminative power, and the combined DTI+SMI model reaching an AUC of only 0.66.

## 4 Discussion

In this study, we systematically compared diffusion-derived metrics from DTI and SMI models across multiple white matter tissue classes in MS, using a large cohort of manually delineated ROIs. Our results demonstrate that both DTI and SMI are sensitive to lesion-related microstructural abnormalities, showing widespread and significant differences across tissue classes. Robust discriminative performance was observed for lesion-NWM and lesion-NAWM comparisons, consistent with pronounced microstructural disruption within lesions relative to surrounding white matter. In contrast, discrimination between NAWM and NWM, as well as between cBHs and T2-lesions, remained limited, suggesting that the microstructural differences between these tissue pairs are relatively subtle. Notably, integrating DTI and SMI parameters resulted in modestly higher AUC values compared with either model alone, supporting the notion that the two models capture complementary aspects of tissue microstructure.

Our findings show that both DTI and SMI metrics are sensitive to microstructural alterations across multiple white matter tissue classes in MS, detecting significant differences between lesions, NAWM, NWM, and lesion subtypes. This pattern suggests that diffusion-based measures capture heterogeneous tissue abnormalities that extend beyond focal lesions. This interpretation is further supported by previous studies reporting widespread DTI abnormalities in NAWM, characterized by increased MD, AD, RD and reduced FA across multiple white matter regions[29, 30]. Importantly, a spatial gradient of FA changes has been described, with more pronounced abnormalities observed in NAWM proximal to lesions and progressively less severe alterations with increasing distance from plaques[31]. Histological evidence of demyelination and axonal transection extending beyond focal lesions further suggests that diffusion abnormalities in NAWM may reflect Wallerian degeneration processes[32-34]. Together, these observations provide a biological explanation for our findings, in which NAWM exhibited significant but relatively subtle diffusion alterations compared with NWM, resulting in limited discriminative performance despite statistically detectable differences.

Building on previous SMI studies in multiple sclerosis, our findings are highly consistent with prior reports showing reduced *p*_2_ and *f*, together with increased 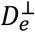, in MS tissue compared with normal white matter. These alterations have been interpreted as reflecting axonal loss, demyelination, and increased microstructural disorganization[35]. Extending beyond these established observations, our ROI-based analysis revealed additional and more nuanced microstructural differences, including significant alterations in 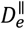, which were not consistently reported in earlier studies. The ability to detect these subtler changes is likely attributable to the large number of carefully manually delineated ROIs and the systematic comparison across multiple lesion types and white matter compartments, highlighting the added sensitivity of detailed regional analyses for characterizing heterogeneous MS pathology[36].

While significant microstructural differences were detected between NAWM and NWM at the group level, the discriminative performance of diffusion-based metrics for this tissue contrast remained limited. This pattern suggests that NAWM abnormalities in MS are present but relatively subtle and spatially heterogeneous, leading to substantial overlap with normal white matter at the individual ROI level[37]. Such observations are consistent with previous diffusion MRI studies, which have reported widespread but diffuse alterations in NAWM, including reduced FA and increased diffusivity (encompassing MD and RD, with more variable findings for AD), indicative of early demyelination and axonal injury rather than focal tissue destruction[38]. Notably, the inconsistency in AD findings may be partly explained by methodological differences; for example, Ahmad et al. did not detect AD abnormalities, likely because their analysis used a single average value per subject for each tissue class, which does not fully exploit the regional heterogeneity across all ROIs[39]. By incorporating data from all individual ROIs, our approach revealed additional subtle but significant AD differences, providing greater insight into the diffuse pathology in NAWM. Importantly, abnormalities in NAWM have also been reported using more advanced diffusion models, with SMT studies demonstrating significant reductions in axonal volume fraction (Vax), further supporting the presence of early axonal pathology within NAWM that may be difficult to robustly discriminate at the regional level [17].

In contrast to lesion-NAWM and lesion-NWM comparisons, differentiation between cBHs and T2-lesions proved more challenging. Although both DTI and SMI metrics revealed statistically significant differences between these lesion subtypes, their overall discriminative performance remained modest. This finding is consistent with previous studies reporting substantial overlap in microstructural properties between cBHs and non-black hole T2 lesions, particularly in early or intermediate disease stages[6, 40]. Pathologically, T2-lesions encompass a heterogeneous spectrum of tissue states, including varying degrees of demyelination, axonal loss, inflammation, and partial remyelination, whereas cBHs are typically associated with more severe axonal destruction [41]. However, incomplete axonal loss within some cBHs and relatively preserved axonal architecture in subsets of T2-lesions may reduce contrast between these lesion types when assessed using diffusion-based metrics alone[42]. Together, these observations suggest that while diffusion models are sensitive to lesion-related microstructural alterations, the biological heterogeneity and partial overlap between cBHs and T2-lesions limit their separability, resulting in only moderate classification performance.

Although SMI-based models consistently achieved slightly higher AUC values than DTI across most tissue contrasts, the overall improvement was modest, indicating that conventional DTI metrics already capture a substantial portion of lesion-related microstructural abnormalities in MS. This observation underscores the continued clinical relevance of DTI, while suggesting that the primary contribution of SMI lies in providing complementary microstructural information rather than replacing tensor-based measures. Importantly, integrating DTI and SMI parameters resulted in consistently higher discriminative performance compared with either model alone, supporting the notion that the two approaches probe partially non-overlapping aspects of tissue microstructure. From a methodological perspective, these findings highlight the value of multi-model diffusion analysis in improving tissue characterization without overreliance on any single metric. Clinically, these modest but consistent gains suggest that advanced diffusion models may enhance, rather than replace, established DTI-based assessments in characterizing white matter pathology in MS.

## 5 Limitation

Several limitations of this study should be acknowledged. First, although many manually delineated ROIs were analyzed, the number of subjects was relatively modest, which may limit patient-level inference and generalizability. Second, our analyses were performed at the ROI level and relied on cross-sectional data, precluding assessment of longitudinal microstructural evolution and causal interpretation of diffusion changes. Third, while significant group-level differences were detected across tissue classes, discriminative performance was limited for NAWM-NWM and cBHs-T2-lesions comparisons, likely reflecting subtle and heterogeneous pathology that overlaps at the individual ROI level. Fourth, although combining DTI and SMI improved classification performance, the observed gains were modest, suggesting that additional advanced diffusion models or multi-shell acquisitions may further enhance tissue characterization. Future studies incorporating longitudinal designs, larger cohorts, and complementary microstructural models will be important to further refine diffusion-based biomarkers of MS pathology

## 6 Conclusion

In summary, this study demonstrates that both DTI and SMI metrics are sensitive to microstructural alterations across a broad spectrum of white matter tissue classes in multiple sclerosis. Using a large set of manually delineated ROIs, we show that diffusion-based measures robustly capture lesion-related abnormalities and more subtle changes extending NAWM. While discriminative performance was high for lesion-white matter contrasts, separability was limited for NAWM versus NWM and between lesion subtypes, reflecting the diffuse and heterogeneous nature of MS pathology. SMI provided modest but consistent improvements over conventional DTI, and their combination yielded the most robust classification performance. Together, these findings support the value of multi-model diffusion MRI for comprehensive characterization of white matter pathology in MS and provide a framework for future studies integrating advanced diffusion models to better capture tissue heterogeneity.

## Data Availability

All data produced in the present study are available upon reasonable request to the authors

## Acknowledgments

We are grateful to our patients and their families, and all the healthy controls who agreed to participate in this study. We thank Mr. Reece Clarke and Mr. Keejin Yoon for their invaluable help in setting up the study and start the recruitment process along with all the MRI technicians of the Vanderbilt University Institute of Imaging Science for assistance with scanning. We would like to express our gratitude to Dr. Harold Moses for his support with recruitment. This manuscript is dedicated to the sweet memory of Mr. Oscar Castr

